# Changes in amygdalar functional connectivity following the water load symptom provocation task (WL-SPT) in youth with functional abdominal pain disorders

**DOI:** 10.1101/2021.05.28.21258026

**Authors:** Natoshia R Cunningham, Hadas Nahman Averbuch, Gregory R. Lee, Christopher King, Robert C. Coghill

**Affiliations:** Department of Family Medicine, Michigan State University; Division of Behavioral Medicine and Clinical Psychology, Cincinnati Children’s Hospital Medical Center; Center for Understanding Pediatric Pain, Cincinnati Children’s Hospital Medical Center; Department of Radiology, Cincinnati Children’s Hospital Medical Center; University of Cincinnati, Department of Radiology; Department of Pediatrics, University of Cincinnati College of Medicine

**Author notes:** Corresponding author: Natoshia R Cunningham, Ph.D., Assistant Professor, Michigan State University, Department of Family Medicine, College of Human Medicine, 628 Secchia Center, 15 Michigan St NE, Grand Rapids, MI, 49503.

**Keywords:** pain, brain, neuroimaging, fMRI, functional connectivity, arterial spin labeling, pediatric, functional abdominal pain, symptom provocation

## Abstract

Pediatric functional abdominal pain disorders (FAPD) are highly prevalent, difficult to diagnose and challenging to treat. The brain mechanisms supporting FAPD remain poorly understood. This investigation examined the neuromechanisms of FAPD during a well-tolerated visceral pain induction task, the water load symptom provocation task (WL-SPT). Youth between the ages of 11 and 17 years participated. Functional connectivity (FC) was examined via the Blood Oxygenation Level Dependent (BOLD) effect using the left and right amygdala (AMY) as seed regions. Relationships of the time courses within these seeds with voxels across the whole brain were evaluated. Arterial spin labeling (ASL) was used to assess regional brain activation by examining cerebral blood flow. Increased FC between the left AMY with regions associated with nociceptive processing (e.g., thalamus), and right AMY FC changes with areas associated with cognitive functioning (dorsolateral prefrontal cortex) and the default mode network (DMN; parietal lobe) were observed in youth with FAPD following the WL-SPT. These changes were related to changes in pain unpleasantness. AMY FC changes post WL-SPT were also related to changes in pain intensity. AMY FC with the DMN in youth with FAPD also differed from healthy controls. Global CBF changes were also noted between FAPD and controls, but no significant differences in grey matter were detected either between groups or during the WL-SPT in youth with FAPD. Findings confirm youth with FAPD have changes in brain connectivity that could support the development of biomarkers to enhance understanding of the mechanisms of pain and treatment response.

## Introduction

Functional abdominal pain disorders (FAPD), chronic and/or recurrent pain conditions, including subtypes such as irritable bowel syndrome (IBS) and functional abdominal pain not otherwise specified [33], are among the most common pain conditions affecting youth [31]. FAPD is thought to result from dysregulated brain-gut interactions [41; 43], yet the neural mechanisms are poorly understood [42].

In adults with IBS, visceral noxious stimulation produces activation in the distributed nociceptive system [12]. Increased activations have been observed in regions associated with *nociceptive processing* (e.g., thalamus, insula [INS], anterior mid cingulate cortex [aMCC], primary [S1] and secondary [S2] somatosensory cortexes) [19; 62], and *emotional arousal* (e.g., the pregenual anterior cingulate cortex [pgACC] and amygdala [AMY]) in individuals with FAPD versus healthy controls [62]. Indeed, FAPD are highly comorbid with anxiety and other emotional problems [11; 15; 17; 64].

Extant pediatric neuroimaging studies suggest structural and functional differences in brain regions associated with attention (e.g., posterior parietal lobe, dorsolateral prefrontal cortex [dlPFC]) and self-reflection (e.g, precuneus, posterior cingulate [PCC]) in adolescents with IBS versus healthy controls [8; 26]. Specifically, cortical thickening in the PCC and thinning in the posterior parietal and dlPFC was observed in youth with IBS versus healthy youth [26]. Lower GMV in regions associated with nociception (e.g., thalamus, aMCC) and attention (dlPFC) was observed in adolescent females with IBS compared to healthy controls [8]. Reduced resting state anti-correlations between regions associated with cognition (e.g., PCC and dlPFC) [26], and reduced resting state functional connectivity (FC) between regions associated with nociception (aMCC) and self-reflection (precuneus) [8] were also observed in those with IBS compared to healthy youth. Less activation in multiple regions associated with attention (e.g., right PCC, right dlPFC) was also found during thermal pain induction via the forearm [25] compared to healthy youth. However, visceral pain was not induced.

In studies of adolescents with IBS that induced non-painful visceral sensations via rectal distention, increased activation in regions associated with nociceptive processing (e.g., insula) and emotional arousal (e.g., pACC) were observed [39]. Reduced hypothalamic FC was observed in brain regions associated with cognition (bilateral prefrontal cortices), memory (hippocampus) and emotion (AMY) [38]. However, there is presently a lack of understanding of the neural mechanisms of pediatric FAPD using non-invasive visceral *pain induction* methods.

The AMY likely plays an important role, given its association with visceral pain and negative affect [45; 47; 54; 57]; yet, there are no studies examining AMY FC in youth with FAPD. We investigated the neural mechanisms of FAPD using the water load symptom provocation task (WL-SPT), a well-tolerated and validated visceral pain induction task [65]. We hypothesized the AMY would demonstrate enhanced resting state FC with other brain regions implicated in pain processing in youth with FAPD following WL-SPT, and that regional brain activation within the distributed nociceptive system would be similarly increased. We further hypothesized that both measures would be related to changes in pain unpleasantness and pain intensity following the WL-SPT. Lastly, we explored baseline differences in resting state FC and regional brain activation between youth with FAPD and healthy controls (HC).

## Methods

### Participants

Participants with FAPD were approached during a visit to a pediatric gastroenterologist at Cincinnati Children’s Hospital Medical Center. Approximately 70% of those approached were interested in participating. Male or female participants were *eligible* if they were between the ages 9-16 years; diagnosed with an FAPD by their pediatric gastroenterologist based on the Rome criteria [23]; English speaking; and had a caregiver willing to provide consent. HC were males and females between 12-17 years old and without chronic pain syndrome who were recruited as part of a separate investigation [46]. All participants were recruited between 2016 and 2019. Participants (FAPD and HC) were *ineligible* if there was evidence of pregnancy; a neurological disorder; MRI counterindications (e.g., orthodontic braces or other metallic implants which obscure or interfere with the MRI); claustrophobia, other significant medical conditions (e.g., Inflammatory Bowel Disease (IBD), diabetes), or a documented developmental delay. Participants were also excluded if they reported current use of psychiatric medication.

In addition to the standard MRI screening processes detailed above, a brief interview to further assess for a fear of enclosed spaces was administered. This interview was adapted from a portion of the *Anxiety Disorder Interview Schedule-Child Report*, a validated semi-structured diagnostic interview to assess for the presence of child psychological disorders [56]. Responses on this module helped to determine eligibility for the study and were used to attempt to reduce instances of unanticipated participant anxiety during the neuroimaging scan.

### Study design

The WL-SPT was only conducted in youth with FAPD. Therefore, neuroimaging changes following the WL-SPT were examined for participants with *FAPD only*, which constituted the primary aims of the study. The exploratory aim utilized HC data obtained from a separate project [46] to compare resting state baseline neuroimaging data in youth with FAPD to those of HC (all scans compared were in the absence of the WL-SPT). The study was approved by the IRB committee of Cincinnati Children’s Hospital Medical Center and written informed consent from a parent/guardian and participant assent were obtained prior to performing study procedures.

FAPD participants completed demographic and psychosocial measures (e.g., anxiety) prior to beginning the WL-SPT task and fMRI scan. Participants then entered the scanner, and scans of brain structure and brain function (BOLD and cerebral blood flow) were performed. Youth provided pain intensity and unpleasantness ratings at the end of each scan. After obtaining the baseline resting-state scans (36 minutes), participants exited the scanner and sat upright in order to ingest water via the WL-SPT (see description below) until they achieved complete fullness (after ∼5 minutes). Following this, they immediately re-entered the scanner and the scans were repeated (21 minutes, post WL-SPT scans). Pain intensity and unpleasantness ratings were also obtained after each scan. In total, the procedures lasted approximately 1.5 hours (∼1 hour total in scanner). Changes in AMY FC and regional brain activation at baseline and following the WL-SPT were investigated.

For the exploratory aim, HC completed the same baseline scans (on the same scanner and the same acquisition parameters) as part of a separate concurrent research investigation by the research team, but did not complete the WL-SPT. Differences in AMY FC and brain activation in the baseline scans of the FAPD youth were then compared to the HC group.

### The water load symptom provocation test (WL-SPT)

The WL-SPT is a validated procedure for children ages 8-16 to create visceral discomfort similar to but less intense than naturally occurring pain sensations experienced by children with FAPD [65] in a safe and non-invasive manner [9] using a standardized protocol [65; 67]. In the validation study, youth reported increased pain symptoms (measures of pain intensity and discomfort which were summed and averaged) from pre to post water load (d = 1.28) as compared to healthy children (d = .89), and these symptoms were less intense than those reported during typical FAPD pain [65].

This procedure requires children to gradually drink water for approximately 5 minutes and up to 15 minutes until they feel “completely full” as assessed by the Fullness Rating Scale [65]. This scale depicted circles which are shaded to represent varying degrees of satiety (empty to completely full), and youth selected the drawing that best depicted their level of fullness. Participants were asked after consuming the water, and if needed after every 5-minutes of drinking thereafter “how much room do you have left?”. They were then asked to point to the imagine depicting their current level of fullness. Water was consumed via a straw from an opaque container containing 1.5 liters of water, which was held by the research assistant so the participant’s perception of satiety were based more on internal versus external cues. Children were instructed to drink water at their own pace. If the participant appeared to be rushing (i.e., gulping), they were instructed not to drink too fast. Children were also asked not eat or drink for 1 hour prior to the study visit.

### Measures

#### Pain assessment

Pain intensity/unpleasantness was assessed via the Visual Analog Scale (VAS). A 15-cm handheld slider scale anchored with the words “no pain intensity/unpleasantness” and “most pain intensity/unpleasantness” [50]. The further the participant moved the slider to the right indicated higher levels of pain symptoms. Numbers on the back of the scale (out of the sight of participants) ranged from 0 to 10. Participants were instructed in the use of these scales via a scripted radio analogy [51]. These scales have been used extensively by this research team in functional imaging studies of pain [13; 34] and they are compatible with use in the MRI scanner. During the fMRI portion of the study, pain intensity and pain unpleasantness were assessed at the end of each scan. A total of 4 intensity and 4 unpleasantness ratings were provided prior to the WL-SPT, and then again after the WL-SPT. These scores were stable within the pre- and post WL-SPT conditions (Figure 1) and therefore scores were explored individually and also averaged within those conditions. These scales have been repeatedly demonstrated (1) reliably separate assessments of intensity and unpleasantness, (2) good internal consistency, and (3) approximate ratio scale measurement accuracy [49].

#### Pain History

Participant socio-demographic factors (e.g., participant age in years, sex, race), was obtained.

#### Functional Disability Inventory (FDI) [63]

A 15-item measure of physical/daily impairment in functioning in the past few days due to health was obtained. The FDI is a widely used, reliable, and valid tool to assess functional disability, and has established cut-offs for mild, moderate, and severe disability [29]. Total scores range from 0-60, with higher scores indicating greater disability.

#### Sample size

No statistical power calculation was conducted prior to the study. The sample size was based on prior research that examined AMY FC in a pediatric chronic pain sample [58], given the limited number of pediatric imaging studies of FAPD.

### Statistical analysis of behavioral data

#### Sociodemographic and clinical data

Statistical analysis of behavioral data was performed using SPSS (version 27). Basic demographic and descriptive data were obtained. Differences between the FAPD and HC groups were then examined using independent samples *t*-tests (age in years, FDI score, average baseline pain unpleasantness and average baseline pain intensity) and chi-square analyses (gender, race). No differences in sociodemographic variables were expected; it was expected that pain-related variables (unpleasantness, intensity, FDI) would be significantly higher in the FAPD group versus the HC group.

#### Pain during the WL-SPT

The average percent change in pain unpleasantness and pain intensity following the WL-SPT was calculated and used in the fMRI analyses.

Two-way ANOVAs were conducted to examine the effect of condition (pre and post WL-SPT) and time (and their interaction) on pain unpleasantness and pain intensity. Significant increases in pain unpleasantness and pain intensity were expected following the WL-SPT in the FAPD youth. Amount of water consumed during the WL-SPT was also recorded and was correlated with changes in pain intensity and pain unpleasantness.

#### Imaging acquisition

All participants were scanned on a 3T Philips Ingenia scanner with a 32 Channel head coil. The total scan time was 1 hour. For FAPD participants, one T1 anatomical scan, two resting-state BOLD scans, then two resting-state pseudo-continuous arterial spin labeling (pCASL) and were completed before the WL-SPT, and then this same series of scans (with BOLDS and pCASL scans in reverse order) was repeated after the WL-SPT when the participant was re-positioned in the scanner. For HC, only the first series of scans was completed. A radiologist reviewed the MRI scans in order to detect incidental findings.

High-resolution T1: multi-echo (4 echoes) weighted images were obtained using the following parameters: TR=10ms, TE=1.8, 3.8, 5.8, 7.8ms, field of view was 256⨯224⨯180mm, voxel size=1⨯1⨯1mm, number of slices=180, flip angle=8 degrees, slice orientation=sagittal, total scan time=4:14 minutes.

BOLD fMRI was performed to examine resting-state connectivity using FOV=240⨯240mm, voxel size=3⨯3mm, slice thickness=4mm, 34 slices, flip angle=90 degrees, TE=35ms, TR=2000ms, number of volumes=150, slice orientation=transverse, dummy scans=2 (data not saved during export from scanner), total scan duration=5:08 minutes.

pCASL was performed to acquire whole-brain cerebral blood flow (CBF) images to examine changes in steady-state regional activation before vs. after WL-SPT in youth with FAPD and also to compare regional activation in youth with FAPD versus HC. Images were acquired using FOV= 240×240, voxel size=2.75×2.75mm, slice thickness=5mm, 22 slices, TR=4.391 seconds, TE=16ms, post label delay=1.8 seconds, label duration=1.65 seconds, number of volumes=66, slice orientation=transverse, dummy scans=0, total scan duration=4:58 minutes.

HC participants were scanned in a parallel study (on the same scanner). The same scan parameters and analysis were used to collect the HC except for a different number of volumes in the BOLD acquisition (193 in HC vs. 150 in FAPD participants) as HC did not undergo the WL- SPT procedure. We used fslroi to truncate the number of volumes to 150 in the HC participants to match the number of volumes in the two groups.

### Image processing and statistical analysis

The FSL (FMRIB’s Software Library, Oxford, UK, version 6.0.0) software package was used for the vast majority of image processing operations and statistical analyses and was augmented by scripts.

T1-weighted images were first bias-corrected using FMRIB’s Automated Segmentation Tool, FAST. The T1-weighted images were brain extracted using the brain extraction tool (BET) and normalized into standard anatomic space (MNI152) using FLIRT (FMRIB’s Linear Image Registration Tool) [27]. Then, images were segmented into the different tissue types and masked with a probability threshold of 0.95 for white matter and CSF.

### BOLD / Connectivity Analyses

For EPI images, we first created a mask of the brain. We then employed slice timing correction using slicetimer and outlier detection using FSL motion outliers routines. Corrupted volumes were identified, and these time points were regressed out of the analysis. In this data set, the mean corrupted volumes were 7.3±4.4 (range of 2-20) in the FAPD group at baseline, 9.0±4.9 (range of 2-24) in the FAPD group after the WL-SPT, and 8.4±5.5 (range of 2-30) in the HC group. Images were then co-registered to the high-resolution T1 structural image and normalized using FMRIB’s Linear Image Registration Tool (FLIRT). A component-based noise correction method (aCompCor) was used to minimize the impact of the physiological, scanner and motion-related artifacts [7]. Based on the CompCor method, a principal component analysis (PCA) was used to characterize the time series data from the white matter and CSF. The PCA of the white matter and CSF was accomplished using the 3DPC module of AFNI (https://afni.nimh.nih.gov/pub/dist/doc/htmldoc/programs/3dpc_sphx.html#ahelp-3dpc) which uses a singular value decomposition. The CompCor method selects the 5 components that accounted for the most variance in the white matter and again for the CSF. These 10 components are included as nuisance regressors in a multiple regression along with motion parameters, consistent with other neuroimaging studies [18; 36; 55]. In addition, 24 motion regressors were included in the GLM design. These regressors were calculated using mcFLIRT and included three regressors for rotation and three for translation. These six regressors were: 1) 1 volume delayed, 2) demeaned and squared, 3) delayed multiplied by the demeaned squared to give a total of 24 motion-related regressors. Denoised residuals were then imported into FEAT and were smoothed with a 5 mm filter and underwent highpass temporal filtering (>0.01 Hz). These images then underwent intensity normalization of each volume in the series. FLIRT was used to register functional images to the high-resolution structural image using boundary-based registration [24], and to the standard space (MNI152) with warp resolution of 10mm.

Registration from high-resolution space to standard space was performed using FNIRT [3]. BOLD images were then placed in standard space by combining the BOLD to structural and the structural to standard transformations.

Time courses of activity were extracted from seed regions. The neuroimaging analyses were pre-planned. Similar to our previous research in pediatric chronic pain samples [44], regions of the right and left AMY were chosen. These seeds were the only seeds that were analyzed. The Juelich atlas was used to create a mask of the seed regions of the right and left AMY using a 50% probability.

The time courses of each seed were separately analyzed using FILM (FMRIB’s Improved Linear Model). First level, fixed-effects analyses were run for each BOLD series to identify voxels that had time courses that were significantly positively and negatively correlated with that of the AMY seed (i.e., seed to whole-brain analysis). Second level fixed effects analyses were used to examine within participants’ effects across imaging series and sessions (pre- and post-WL-SPT). Finally, third level random effects analyses (FMRIB’s Local Analysis of Mixed Effects, FLAME 1+2) were employed to identify mean effect of the WL-SPT by comparing differences in functional connectivity before and after the WL-SPT. Demeaned values of percent changes in pain unpleasantness and pain intensity after the WL-SPT were also included in the model as a separate covariate of interest (changes in pain intensity were orthogonalized to changes in pain unpleasantness). Clusters of connectivity were identified using a threshold of Z>2.3. Cluster significance of p<0.05 was estimated according to Gaussian random field theory [71].

In order to identify alterations in amygdalar connectivity in participants with FAPD, we compared the baseline resting-state functional connectivity (before WL-SPT) of the patient group with HC data collected in parallel (2016-2017) for another study [46]. In this analysis, third level random effects analyses (FMRIB’s Local Analysis of Mixed Effects, FLAME 1+2) were employed to identify differences in resting state FC between participants with FAPD and HC. Clusters of connectivity were identified using a threshold of Z>2.3. Cluster significance of p<0.05 was estimated according to Gaussian random field theory [71].

### Preprocessing of pCASL images into CBF images

CBF was quantified in units of (ml of blood)/min/(100 g of tissue) using in-house software that implements an established quantitative model [66]. This process yielded a single, fully quantified CBF volume (mL/100g/min) from each 4D series of pCASL images. To accomplish this, the 4D pCASL series was first motion corrected. Next, a perfusion-weighted series was generated by subtracting tag and control images. Due to the relatively small signal changes observed with ASL, even after motion correction, some motion-related outliers in the “control - tag” subtracted images may be present. Outliers were detected in two stages and excluded from the time series average. In stage one, any (control-tag) points corresponding to a mean signal change far outside of a physiologically plausible CBF-related range were discarded. We chose to use a mean signal change of > 3% relative to the control image for this metric (typical CBF-related signal changes are <2% in GM and much lower in WM, so a 3% signal change over the whole brain is physiologically implausible). In the second stage FSL’s fsl_motion_outliers.sh script was used to compare the root-mean-squared intensity difference between a reference volume and each time point. Outliers were identified as those falling more than 1.5 times the interquartile range above the 75th percentile. A minor modification was made to the script to use the time series average subtraction as the reference for comparison rather than choosing a single individual subtraction as the reference point. A total of 33 pairs were collected. The mean numbers of pairs kept in the primary analysis were 30.6±2.6 and 30.6±2.9 (before and after WL-SPT, respectively).

A gray matter T1 value of 1.3 s was assumed and sensitivity of the quantitative CBF value to tissue T1 was reduced by using a post-labeling delay, which is longer than the transit time from the labeling plane to the imaging slice [1]. Background suppression of static tissue was used to improve the stability of the ASL time series [2]. In order to estimate the equilibrium magnetization, M0, for use in the quantitative model, a separate short reference scan without background suppression, but otherwise identical parameters were acquired. The magnetization density, M0, in the blood is estimated based on the measured signal amplitude from this reference scan in white matter (due to its short T1, WM is nearly fully relaxed at our TR of 4.4s). The M0 in white matter was used to approximate M0 of the arterial blood by multiplying by a factor R = 0.87 / 0.73 that reflects the relative spin density of blood vs. white matter [22].

CBF images were smoothed with an 8mm FWHM Gaussian filter. The four image files (two pre-WL-SPT and two post-WL-SPT) were concatenated in the primary analysis to produce one single output series with four volumes. Additional image processing was accomplished in FEAT (FMRI Expert Analysis Tool) Version 6.0.0. The CBF series was motion-corrected and intensity normalized to minimize the impact of fluctuations in global CBF. FLIRT was used to register CBF images to the high-resolution structural image using boundary-based registration [24]. Registration of the high-resolution structural volume to standard space (MNI152) was performed using FNIRT nonlinear registration with a warp resolution of 10mm [3]. CBF images were then placed in standard space by combining the CBF to structural and the structural to standard transformations. A first-level fixed-effects analysis was executed using FEAT to identify within participant effects. The differences in CBF between pre- vs post-WL-SPT were identified. The higher-level analysis was carried out using FLAME (FMRIB’s Local Analysis of Mixed Effects) stage 1 [6; 69]. A second-level random effects analysis was executed within FEAT to identify between participant effects related to WL-SPT. Demeaned values of percent changes in pain unpleasantness and pain intensity after the WL-SPT were also included in the model as a separate covariate of interest. For these analyses, changes in pain intensity were orthogonalized to changes in pain unpleasantness. Clusters were identified using a threshold of Z>2.3 and a (corrected) cluster significance threshold of p=0.05 [70].

In order to identify differences in regional brain activation among participants with FAPD as compared to HC, we also compared the resting-state baseline pCASL data (before WL- SPT) of the FAPD group to the HC group. In this analysis, non-parametric general linear model-based analyses using permutation inference to identify differences in resting state pCASL data between participants with FAPD and HC (FSL randomise, 5000 permutations)[68]. Clusters of connectivity were identified using threshold free cluster enhancement (TFCE) [59]. Cluster significance was established at p<0.05, with family wise error correction to control for multiple comparisons across voxels.

## Results

### Descriptive data

A total of twenty-seven youth with FAPD were recruited the current study. One FAPD participant was found to have an arachnoid cyst and another FAPD participant was subsequently diagnosed with IBD and thus were excluded from study analyses. No other incidental findings were reported in either the FAPD or HC group. No participants withdrew during the scanning phase of the study. A total of 25 FAPD participants (13 females, 12 males; mean age of 13.4 years; SD: 2.2) and 20 HC (17 females, 3 males; mean age of 14.7 years; SD: 1.8) were included in the study analysis. Seventeen (68%) FAPD participants had IBS and 8 (32%) had FAPD- NOS. Information on participants’ demographic and psychosocial characteristics are presented in Table 1. FAPD and HC groups differed in sex (*X*^*2*^ = 5.45, *p* = .020) and age (*t*(43) = 2.18, *p* < .035), with more females and slightly older youth in the HC group. As expected, the groups differed in baseline FDI scores (*t*(43) = 6.00, *p* < .001), baseline pain unpleasantness during resting-state scans (*t*(43) = 4.47, *p* < .001), and baseline pain intensity during resting-state scans (*t*(43) = 4.29, *p* < .001).

### Changes in pain symptoms following the WL-SPT in youth with FAPD

After the WL-SPT, pain unpleasantness increased an average of 49% across all scans from 1.72 to 2.56. In addition, pain intensity increased 29%, from 1.79 to 2.31. Almost all FAPD participants experienced increases in pain unpleasantness (96%) and pain intensity (84%) following the WL-SPT. Pain unpleasantness decreased in 1 participant, and pain intensity was unchanged in 1 participant and decreased in 3 participants following the WL-SPT.

On average 461.53 ml (sd = 237.76) of water was consumed during the WL-SPT (range 177.44 - 957.93). Water consumption was not significantly correlated with average change in pain unpleasantness, but there was a trending association whereby more water consumption was related to increased pain unpleasantness (r = .37, p = 0.075). Water consumption was not correlated with changes in pain intensity (r = 0.04, p = 0.855).

The two-way ANOVAs revealed significant differences by condition (pre or post WL- SPT) on pain unpleasantness (F (1, 24) = 20.241, p < .001; main effect p < 0.001) and pain intensity (F (1, 24) = 7.215, p = .013; main effect p = 0.013) (Figure 1). There was no significant differences by time, nor were there time by condition interactions (all p’s > 0.05).

### AMY FC before and after the WL-SPT in youth with FAPD

Data from all 25 participants with FAPD were included in these analyses. Lower FC between the left AMY and areas that are classified as white matter were found following the WL-SPT as compared to baseline. No changes in resting state FC were detected for the right AMY between baseline and following the WL-SPT.

Increased *pain unpleasantness* following the WL-SPT were related to increased FC between the left AMY and the cerebellum, right thalamus & right caudate in youth with FAPD (Figure 2). Changes in pain unpleasantness following the WL-SPT were also related to increased FC between the right AMY and dorsolateral prefrontal cortex and a reduction in FC between the right AMY and the left parietal and central opercular cortex, superior parietal lobe, postcentral gyrus, bilateral SMA, precentral gyrus, & superior frontal gyrus in youth with FAPD.

Changes in *pain intensity* following the WL-SPT were related to increased FC between the left AMY and the OFC and subgenual ACC in youth with FAPD (Figure 3). Changes in pain intensity following the WL-SPT were also related to increased FC between the right AMY and the bilateral occipital cortex, right cerebellum, subgenual ACC, right middle frontal gyrus and left caudate.

### Regional brain activation before and after the WL-SPT in youth with FAPD

Four participants were excluded from the pCASL analysis due to unsuccessful labeling of one or more blood vessel(s), which led to absent signal in the corresponding vascular territories of the brain. Thus, the remaining data from 21 participants with FAPD were used for the pCASL analyses. The results revealed no statistically significant differences in CBF before versus after the WL-SPT in participants with FAPD. In addition, there was no significant association between regional brain activation with changes in pain symptoms (unpleasantness and intensity) following the WL-SPT.

### AMY FC differences in youth FAPD versus HC at rest

Twenty participants with FAPD were compared with 18 HC. Participants with FAPD had *increased* FC between the left AMY and the left precuneus, right superior and middle frontal gyrus, right inferior parietal lobule and the bilateral lateral occipital cortex (Figure 4; Table 2). There was no greater FC connectivity between the left AMY in HC versus FAPD participants.

Participants with FAPD had *increased* FC between the right AMY and the bilateral PCC, precuneus & left inferior parietal cortex. HC had *increased* FC between the right AMY and the bilateral occipital pole & lateral occipital cortex.

### Activation differences in youth FAPD versus HC at rest

Twenty participants with FAPD were compared with 18 HC. One additional participant was lost from the FAPD group due to incomplete coverage of the brain during acquisition and two additional participants were lost from the HC group due to incomplete tagging of blood vessels.

Global cerebral blood flow differed significantly between FAPD patients and HC (45.20±12.46 ml/100g/min vs. 53.64±8.20 ml/100g/min, respectively, t(1,36)=2.467, p=0.019). Consistent with these absolute changes in global cerebral blood flow, regional differences in relative CBF were evident in the white matter. White matter CBF with the FAPD group exhibited greater relative CBF in comparison to HC. Conversely, large amounts of grey matter near the top of the brain exhibited reduced activation in the FAPD group relative to HC. This was largely due to artifactual reductions in CBF in the FAPD group arising from tagging issues in the top slices of several participants. No significant changes in regional activation in grey matter areas were noted, although trends for reduced relative CBF in the FAPD group were detected in the right posterior insula (p<0.08), right hippocampus (p<0.07), and left primary somatosensory cortex (p<0.07).

## Discussion

Following a validated symptom provocation task (the WL-SPT), substantive differences in functional connectivity in youth with FAPD were observed that were related to increases in pain unpleasantness and pain unpleasantness. Further, the brains of youth with FAPD at rest operate differently when compared to HCs. Namely, functional connectivity patterns of the amygdala (and to a lesser extent regional brain activation) in youth with FAPD significantly differ from healthy children, which both supports [62] and expands upon prior research. These findings provide a foundation that may lead to the discovery of biomarkers to aid in better identifying FAPDs.

### The role of the AMY in pain processing

In youth with chronic pain, the AMY is thought to play an important role in aspects of nociceptive processing [57; 61], including pain attention and anticipation of pain [40; 60]. Prior research has shown the AMY was activated during visceral pain in adults with IBS compared to healthy controls [62] and changes in hypothalamus FC with the AMY were observed in youth with IBS versus healthy youth undergoing non-painful rectal distention [38]. AMY FC has also changed following treatment for other pediatric chronic pain conditions, such as CRPS [58] and migraine [45]. Interestingly, the AMY is also associated with negative affect, hyperarousal, and stress, and FAPD are highly comorbid with anxiety and other emotional problems [11; 15; 17; 64], particularly in more persistent and severe cases [16].

### Linking neuroimaging data to subjective reports of pain

Few studies in general have linked subjective reports of pain to brain imaging data in pediatric FAPD [26]. Our findings revealed pain unpleasantness in particular may be an important outcome of the WL-SPT in youth with FAPD as it increased by a significant margin (49%) and was found to be related to changes in AMY FC. Changes in pain unpleasantness following the WL-SPT were related to 1) increased FC between the left AMY and the cerebellum, right thalamus (associated with nociceptive processing) & right caudate, 2) increased FC between the right AMY and dorsolateral prefrontal cortex (associated with cognition) and 3) a reduction in FC between the right AMY and the left parietal and central operculum cortex, superior parietal lobe, postcentral gyrus, bilateral SMA, precentral gyrus, & superior frontal gyrus. These findings support prior research suggesting the AMY is activated during attention to an unpleasant noxious stimulus [37]. Pain intensity following the WL-SPT was also related to increased FC between the left and right AMY with various brain regions potentially implicated in pain (e.g., the OFC, right cerebellum, ACC, right middle frontal gyrus and left caudate).

### FAPD versus HC

There were also multiple differences observed in resting-state FC patterns in youth with FAPD versus HC, with FAPD participants generally showing enhanced AMY FC as compared to HC. In our study, participants with FAPD had *increased FC* 1) between the left AMY and the left precuneus, right superior and middle frontal gyrus, right inferior parietal lobule and the bilateral lateral occipital cortex, and 2) between the right AMY and the left inferior parietal cortex, bilateral PCC, and the precuneus, when compared with HC. Of note, the PCC and precuneus regions are considered components of the default mode network (DMN). The DMN is active when the mind is wandering (i.e., not engaged in a specific task) [4; 5; 52], and may be engaged when youth with chronic pain experience self-referential thoughts about their pain. In a prior a somatic pain induction study of adolescents with and without chronic pain, significant activations were noted in the DMN, and youth with chronic pain also demonstrated increased activation in the PCC compared to controls [28]. Evidence of *increased* AMY FC with DMN regions in the current study is in line with other research showing increased activity in similar brain regions in youth with chronic pain [58]. Therefore, these findings may represent altered affective-cognitive processing in youth with FAPD. Future research directly comparing brain imaging data of youth with FAPD to HC *during* the WL-SPT would also be informative.

### Using a validated symptom provocation task

Our investigation was also the first to use validated visceral pain induction technique during neuroimaging in youth with FAPD. This task evoked mild visceral pain sensations similar to but not as severe as a typical FAPD pain flare [65], which is critical for feasibility in pediatric populations. Prior studies imaged FAPD participants at rest [26], or used somatic (versus visceral) pain provocation [8; 25], which is conceptually discordant to the visceral pain experienced during an FAPD flare. Of the limited studies that have used invasive visceral symptom provocation methods (e.g., rectal distention) [38; 39; 62], they by design did not evoke pain symptoms in pediatric samples [38; 39], limiting the interpretation of study findings. Furthermore, rectal distention is invasive and aversive, and can skew recruitment. Using a visceral pain induction task mimics a true FAPD pain episode and may provide better understanding on the mechanisms related to FAPD.

The WL-SPT is a safe, effective, and non-invasive strategy for inducing visceral sensations similar to true FAPD pain. A relatively high participant agreement rate (70%) in the current study supports its feasibility and acceptability. Not only did the WL-SPT reliably increase pain unpleasantness (and to a lesser degree pain intensity), but these changes were associated with changes observed in AMY FC. *Prior to the current investigation, the WL-SPT had never before been used during neuroimaging to study pediatric FAPD pain* and had infrequently been used in adult studies [32]. Clearly, this task is an appropriate method of visceral pain induction in pediatric neuroimaging research.

### Regional and global CBF during functional abdominal pain

Multiple methods (ASL and BOLD fMRI) were also used to attempt to capture different dynamic aspects of pain. ASL is fully quantitative and is considered optimal for imaging relatively slow or steady-state phenomena [20], whereas BOLD FC is useful for capturing rapidly fluctuations in brain activity. We have previously used ASL to examine acute heat pain in healthy adults in studies of meditation [72; 73] as well as effects of CBT in youth with migraine [45]. Of note, global CBF was significantly lower in FAPD patients than in HC. Global CBF reductions are frequently associated with hyperventilation and the concomitant reduction of pCO_2_. Given youth with FAPD have high levels of anxiety and that anxiety is associated with hyperventilation, it is possible that different resting ventilation rates between groups accounted for these differences in global CBF [14; 30; 53]. Despite these global differences in CBF, there were trending differences in regional normalized CBF between youth with FAPD versus healthy youth that occurred in brain areas associated with pain.

Unexpectedly, there were no differences in regional brain activation as assessed via normalized CBF (even when examined in relation to pain unpleasantness/intensity) after the WL-SPT in participants with FAPD. Data from 4 participants was lost due to tagging artifacts, but the final group size (21) was similar to those of our previous studies. However, the presence of significant pain at rest in the participants with FAPD may have reduced the ability to detect additional activation evoked by the WL-SPT.

### Strengths, limitations, and future directions

Much of the neuroimaging research in FAPD pain has been limited to adults who may have experienced chronic pain for many years. Therefore, these findings are important, like other pediatric chronic pain studies in CRPS [58], and IBS [8; 26], as they suggest childhood pain may re-wire neural circuitry and contribute to fundamental differences in pain processing compared to healthy youth. These findings are also important given the increased potential for neuroplasticity in pediatric populations. Our study has advanced understanding of pediatric FAPD as the sample included some younger pediatric FAPD youths and was not limited to a specific FAPD subtype. Prior studies of pediatric FAPD used adolescent participants with the IBS subtype only [26; 38; 39]. Our sample is truly representative of the youth with FAPD who present to pediatric GI clinics for care. Limitations include a relatively small sample, HC had differences in sociodemographic characteristics and was obtained from a different study, and lack of control for FAPD subtypes.

## Data Availability

Data is available upon request.

## Summary

Overall, findings increase understanding of the pain experience in youth with FAPD, and the neural mechanisms underlying chronic pain. Cumulatively, these findings support the notion that the impact of chronic pain is broad-ranging [10; 48] and may affect multiple networks [35]. Specifically, changes observed in AMY FC with areas associated with cognitive functioning and nociceptive processing may lead to more mechanistically derived biomarkers for FAPD pain. Such biomarkers would represent a transformative advancement as these conditions are traditionally considered disorders of exclusion, and commonly diagnosed only after a lengthy, and potentially risky and expensive medical work-up [21]. There are limited evidence-based treatments currently available for FAPD; therefore, such findings also have the potential to inform the development and testing of personalized and effective treatment approaches.

## Acknowledgements

We thank the study staff in the Division of Behavioral Medicine and Clinical Psychology at Cincinnati Children’s Hospital Medical Center who contributed to the successful execution of this research, including Erin Moorman and Christine Le. We also thank Samantha Ely who assisted with the analysis and presentation of data. We also owe our utmost thanks to the children who participated in the study. The study was funded in part by an NIH/NCCIH K23 award, AT009458 (to the first author NRC) in addition to startup funds (for RCC and CDK) and a Discovery Award from Cincinnati Children’s Hospital (for CDK). The authors declare no conflicts of interest.

## Figure legends

**Figure 1. Pain intensity and pain unpleasantness ratings before and after the WL-SPT in youth with FAPD**. Four pain intensity and pain unpleasantness ratings were provided prior to the WL-SPT, and then again after the WL-SPT (after each respective scan). Two-way ANOVAs revealed significant differences by condition (pre versus post WL-SPT) on pain unpleasantness (omnibus, F (1, 24) = 20.241, p < .001; main effect, p < 0.001) and pain intensity (omnibus, F (1, 24) = 7.215, p = .013; main effect, p = 0.013). There weas no significant differences by time, nor was there a time by condition interaction (all p’s > 0.05). Thus, these scores were stable within the pre- and post WL-SPT conditions WL-SPT- water load symptom provocation task; FAPD- functional abdominal pain disorders

**Figure 2. Differences in AMY FC in relation to changes in pain unpleasantness after the WL-SPT in youth with FAPD**. In youth with FAPD, changes in pain unpleasantness following the WL-SPT were related to 1) increased FC between the left AMY and the cerebellum, right thalamus and right caudate in youth with FAPD, 2) increased FC between the right AMY and dorsolateral prefrontal cortex, and 3) a reduction in FC between the right AMY and the left parietal and central operculum cortex, superior parietal lobe, postcentral gyrus, bilateral SMA, precentral gyrus, and superior frontal gyrus.

AMY FC- Amygdalar functional connectivity; WL-SPT- water load symptom provocation task; FAPD- functional abdominal pain disorders; SMA- supplementary motor area.

**Figure 3. Differences in AMY FC in relation to changes in pain intensity after the WL- SPT**. In youth with FAPD, changes in pain intensity following the WL-SPT were related to 1) increased FC between the left AMY and the OFC, and 2) increased FC between the right AMY and the bilateral occipital cortex, right cerebellum, ACC, right middle frontal gyrus and left caudate.

AMY FC- Amygdalar functional connectivity; WL-SPT- water load symptom provocation task; AMY- Amygdala; FAPD- functional abdominal pain disorders; OFC -orbitofrontal cortex; ACC– anterior cingulate cortex.

**Figure 4. Differences in resting-state AMY FC in youth with FAPD as compared to HC**. Participants with FAPD demonstrated increased resting-state FC between the left AMY and the left precuneus, right superior and middle frontal gyrus, right inferior parietal lobule and the bilateral lateral occipital cortex. There was no greater AMY FC in HC versus FAPD participants. Participants with FAPD also had increased FC between the right AMY and the bilateral PCC, precuneus and left inferior parietal cortex. HC had increased FC between the right AMY and the bilateral occipital pole and lateral occipital cortex.

AMY FC- Amygdalar functional connectivity; WL-SPT- water load symptom provocation task; FAPD- functional abdominal pain disorders; HC- healthy controls.

## Summary

Youth with FAPD have altered amygdalar connectivity following a visceral pain induction task that was associated with changes in pain unpleasantness and pain intensity. Differences in amygdalar connectivity in youth with FAPD were also observed at rest when compared with healthy youth.

## Notes

### Competing Interest Statement

The authors have declared no competing interest.

### Funding Statement

The study was funded in part by an NIH NCCIH K23 award to the first author NRC in addition to startup funds for RCC and CDK and a Discovery Award from Cincinnati Childrens Hospital for CDK

### Author Declarations

Cincinnati Children's Hospital Medical Center

## References

[1] Alsop DC, Detre JA. Multisection cerebral blood flow MR imaging with continuous arterial spin labeling. Radiology 1998;208:410–416.

[2] Alsop DC, Detre JA, Golay X, Gunther M, Hendrikse J, Hernandez-Garcia L, Lu H, MacIntosh BJ, Parkes LM, Smits M, van Osch MJ, Wang DJ, Wong EC, Zaharchuk G. Recommended implementation of arterial spin-labeled perfusion MRI for clinical applications: A consensus of the ISMRM perfusion study group and the European consortium for ASL in dementia. Magn Reson Med 2015;73:102–116.

[3] Andersson JLRJ, M; Smith, S. Non-linear optimisation. FMRIB technical report 2007.

[4] Baliki MN, Chialvo DR, Geha PY, Levy RM, Harden RN, Parrish TB, Apkarian AV. Chronic pain and the emotional brain: specific brain activity associated with spontaneous fluctuations of intensity of chronic back pain. J Neurosci 2006;26:12165–12173.

[5] Baliki MN, Geha PY, Apkarian AV, Chialvo DR. Beyond feeling: chronic pain hurts the brain, disrupting the default-mode network dynamics. J Neurosci 2008;28:1398–1403.

[6] Beckmann CF, Jenkinson M, Smith SM. General multilevel linear modeling for group analysis in FMRI. Neuroimage 2003;20:1052–1063.

[7] Behzadi Y, Restom K, Liau J, Liu TT. A component based noise correction method (CompCor) for BOLD and perfusion based fMRI. Neuroimage 2007;37:90–101.

[8] Bhatt RR, Gupta A, Labus JS, Zeltzer LK, Tsao JC, Shulman RJ, Tillisch K. Altered brain structure and functional connectivity and its relation to pain perception in girls with irritable bowel syndrome. Psychosom Med 2019;81:146.

[9] Birnie KA, Caes L, Wilson AC, Williams SE, Chambers CT. A practical guide and perspectives on the use of experimental pain modalities with children and adolescents. Pain Manag 2014;4:97–111.

[10] Bushnell MC, Čeko M, Low LA. Cognitive and emotional control of pain and its disruption in chronic pain. Nat Rev Neurosci 2013;14:502–511.

[11] Campo JV, Bridge J, Ehmann M, Altman S, Lucas A, Birmaher B, Di Lorenzo C, Iyengar S, Brent DA. Recurrent abdominal pain, anxiety, and depression in primary care. JAMA Pediatr 2004;113:817–824.

[12] Coghill RC. The Distributed Nociceptive System: A Framework for Understanding Pain. Trends Neurosci 2020.

[13] Coghill RC, McHaffie JG, Yen YF. Neural correlates of interindividual differences in the subjective experience of pain. Proc Natl Acad Sci USA 2003;100:8538–8542.

[14] Coghill RC, Sang CN, Berman KF, Bennett GJ, Iadarola MJ. Global cerebral blood flow decreases during pain. J Cereb Blood Flow Metab 1998;18:141–147.

[15] Cunningham NR, Cohen MB, Farrell MK, Mezoff AG, Lynch-Jordan A, Kashikar-Zuck S. Concordant parent-child reports of anxiety predict impairment in youth with functional abdominal pain. J Pediatr Gastroenterol Nutr 2014;60:312–317.

[16] Cunningham NR, Jagpal A, Peugh J, Cohen MB, Farrell MK, Mezoff AG, Lynch AM, Kashikar-Zuck S. Using a simple risk categorization to predict outcomes in pediatric functional abdominal pain at 6 months J Pediatr Gastroenterol Nutr 2017;64:685–690.

[17] Cunningham NR, Lynch-Jordan A, Mezoff AG, Farrell MK, Cohen MB, Kashikar-Zuck S. Importance of addressing anxiety in youth with functional abdominal pain: suggested guidelines for physicians. J Pediatr Gastroenterol Nutr 2013;56:469–474.

[18] Davey J, Thompson HE, Hallam G, Karapanagiotidis T, Murphy C, De Caso I, Krieger-Redwood K, Bernhardt BC, Smallwood J, Jefferies E. Exploring the role of the posterior middle temporal gyrus in semantic cognition: Integration of anterior temporal lobe with executive processes. Neuroimage 2016;137:165–177.

[19] Derbyshire SW. Visceral afferent pathways and functional brain imaging. Sci World J 2003;3:1065–1080.

[20] Detre JA, Rao H, Wang DJ, Chen YF, Wang Z. Applications of arterial spin labeled MRI in the brain. J Magn Reson Imaging 2012;35:1026–1037.

[21] Dhroove G, Chogle A, Saps M. A Million-dollar Work-up for Abdominal Pain: Is It Worth It? J Pediatr Gastroenterol Nutr 2010;51:579–583.

[22] Donahue MJ, Lu H, Jones CK, Edden RA, Pekar JJ, van Zijl PC. Theoretical and experimental investigation of the VASO contrast mechanism. Magn Reson Med 2006;56:1261–1273.

[23] Drossman DA, Dumitrascu DL. Rome III: New standard for functional gastrointestinal disorders. J Gastrointestin Liver Dis 2006;15:237–241.

[24] Greve DN, Fischl B. Accurate and robust brain image alignment using boundary-based registration. Neuroimage 2009;48:63–72.

[25] Huang JS, Terrones L, Simmons AN, Kaye W, Strigo I. A pilot study of fMRI responses to somatic pain stimuli in youth with functional and inflammatory gastrointestinal disease. J Pediatr Gastroenterol Nutr 2016;63:500.

[26] Hubbard CS, Becerra L, Heinz N, Ludwick A, Rasooly T, Wu R, Johnson A, Schechter N, Borsook D, Nurko S. Abdominal Pain, the Adolescent and Altered Brain Structure and Function. PLoS ONE 2016;11:e0156545.

[27] Jenkinson M, Bannister P, Brady M, Smith S. Improved optimization for the robust and accurate linear registration and motion correction of brain images. Neuroimage 2002;17:825–841.

[28] Jones SA, Cooke HE, Wilson AC, Nagel BJ, Holley AL. A pilot study examining neural response to pain in adolescents with and without chronic pain. Front Neurol 2020;10:1403.

[29] Kashikar-Zuck S, Flowers S, Claar R, Guite J, Logan D, Lynch-Jordan A, Palermo T, Wilson A. Clinical utility and validity of the Functional Disability Inventory among a multicenter sample of youth with chronic pain. Pain 2011;152:1600–1607.

[30] Kety SS, Schmidt CF. The effects of active and passive hyperventilation on cerebral blood flow, cerebral oxygen consumption, cardiac output, and blood pressure of normal young men. J Clin Invest 1946;25:107–119.

[31] King S, Chambers CT, Huguet A, MacNevin RC, McGrath PJ, Parker L, MacDonald AJ. The epidemiology of chronic pain in children and adolescents revisited: A systematic review. Pain 2011;152:2729–2738.

[32] Kleinhans NM, Yang CC, Strachan ED, Buchwald DS, Maravilla KR. Alterations in connectivity on functional magnetic resonance imaging with provocation of lower urinary tract symptoms: a MAPP research network feasibility study of urological chronic pelvic pain syndromes. J Urol 2016;195:639–645.

[33] Koppen IJ, Nurko S, Saps M, Di Lorenzo C, Benninga MA. The pediatric Rome IV criteria: what’s new? Expert Rev Gastroenterol Hepatol 2017;11:193–201.

[34] Koyama T, McHaffie JG, Laurienti PJ, Coghill RC. The single-epoch fMRI design: validation of a simplified paradigm for the collection of subjective ratings. Neuroimage 2003;19:976–987.

[35] Kucyi A, Davis KD. The dynamic pain connectome. Trends Neurosci 2014.

[36] Kucyi A, Moayedi M, Weissman-Fogel I, Goldberg MB, Freeman BV, Tenenbaum HC, Davis KD. Enhanced medial prefrontal-default mode network functional connectivity in chronic pain and its association with pain rumination. J Neurosci 2014;34:3969–3975.

[37] Kulkarni B, Bentley D, Elliott R, Youell P, Watson A, Derbyshire S, Frackowiak R, Friston K, Jones A. Attention to pain localization and unpleasantness discriminates the functions of the medial and lateral pain systems. Eur J Neurosci 2005;21:3133–3142.

[38] Liu X, Li S-J, Shaker R, Silverman A, Kern M, Ward BD, Li W, Xu Z, Chelimsky G, Sood MR. Reduced functional connectivity between the hypothalamus and high-order cortical regions in adolescent patients with irritable bowel syndrome. J Pediatr Gastroenterol Nutr 2017;65:516.

[39] Liu X, Silverman A, Kern M, Ward BD, Li SJ, Shaker R, Sood MR. Excessive coupling of the salience network with intrinsic neurocognitive brain networks during rectal distension in adolescents with irritable bowel syndrome: a preliminary report. Neurogastroenterol Motil 2016;28:43–53.

[40] Lobanov OV, Quevedo AS, Hadsel MS, Kraft RA, Coghill RC. Frontoparietal mechanisms supporting attention to location and intensity of painful stimuli. Pain 2013;154:1758–1768.

[41] Lydiard RB. Irritable bowel syndrome, anxiety, and depression: What are the links? J Clin Psychiatr 2001;62:38–45.

[42] Mayer EA, Aziz Q, Coen S, Kern M, Labus J, Lane R, Kuo B, Naliboff B, Tracey I. Brain imaging approaches to the study of functional GI disorders: a Rome working team report. Neurogastroenterol Motil 2009;21:579–596.

[43] Mayer EA, Tillisch K. The brain-gut axis in abdominal pain syndromes. Annu Rev Med 2011;62:381–396.

[44] Nahman-Averbuch H, Schneider VJ, 2nd, Chamberlin LA, Kroon Van Diest AM, Peugh JL, Lee GR, Radhakrishnan R, Hershey AD, King CD, Coghill RC, Powers SW. Alterations in Brain Function After Cognitive Behavioral Therapy for Migraine in Children and Adolescents. Headache 2020.

[45] Nahman-Averbuch H, Schneider VJ, Chamberlin LA, Van Diest Amk, Peugh JL, Lee GR, Radhakrishnan R, Hershey AD, Powers SW, Coghill RC. Identification of neural and psychophysical predictors of headache reduction after cognitive behavioral therapy in adolescents with migraine. Pain 2021;162:372–381.

[46] Nahman-Averbuch H, Schneider VJ, Lee GR, Peugh JL, Hershey AD, Powers SW, Coghill RC, King CD. Neural mechansims of migrain in adolescents: relationships with sleep. under review.

[47] Neugebauer V, Li W, Bird GC, Han JS. The amygdala and persistent pain. Neuroscientist 2004;10:221–234.

[48] Peyron R, Laurent B, Garcia-Larrea L. Functional imaging of brain responses to pain. A review and meta-analysis (2000). Clin Neurophysiol 2000;30:263–288.

[49] Price DD. Psychological and Neural Mechanisms of the Affective Dimension of Pain. Science 2000;288:1769–1772.

[50] Price DD, Bush FM, Long S, Harkins SW. A comparison of pain measurement characteristics of mechanical visual analogue and simple numerical rating scales. Pain 1994;56:217–226.

[51] Price DD, McHaffie GJ, Larson MA. Spatial summation of heat-induced pain: influence of stimulus area and spatial separation of stimuli on perceived pain sensation intensity and unpleasantness. J Neurophysiol 1989;62:1270–1279.

[52] Raichle ME, MacLeod AM, Snyder AZ, Powers WJ, Gusnard DA, Shulman GL. A default mode of brain function. Proc Natl Acad Sci USA 2001;98:676–682.

[53] Ramsay S, Murphy K, Shea S, Friston K, Lammertsma A, Clark J, Adams L, Guz A, Frackowiak R. Changes in global cerebral blood flow in humans: effect on regional cerebral blood flow during a neural activation task. J Physiol 1993;471:521–534.

[54] Sadler KE, McQuaid NA, Cox AC, Behun MN, Trouten AM, Kolber BJ. Divergent functions of the left and right central amygdala in visceral nociception. Pain 2017;158:747.

[55] Shpaner M, Kelly C, Lieberman G, Perelman H, Davis M, Keefe FJ, Naylor MR. Unlearning chronic pain: A randomized controlled trial to investigate changes in intrinsic brain connectivity following Cognitive Behavioral Therapy. Neuroimage Clin 2014;5:365–376.

[56] Silverman WK, Albano A. The Anxiety Disorders Interview Schedule for Children for DSM-IV (child and parent versions). San Antonio, TX: Psychological Corporation, 1996.

[57] Simons LE, Moulton EA, Linnman C, Carpino E, Becerra L, Borsook D. The human amygdala and pain: evidence from neuroimaging. Human Brain Mapp 2014;35:527–538.

[58] Simons LE, Pielech M, Erpelding N, Linnman C, Moulton E, Sava S, Lebel A, Serrano P, Sethna N, Berde C, Becerra L, Borsook D. The responsive amygdala: treatment-induced alterations in functional connectivity in pediatric complex regional pain syndrome. Pain 2014;155:1727–1742.

[59] Smith SM, Nichols TE. Threshold-free cluster enhancement: addressing problems of smoothing, threshold dependence and localisation in cluster inference. Neuroimage 2009;44:83–98.

[60] Strigo IA, Simmons AN, Matthews SC, Arthur D. The relationship between amygdala activation and passive exposure time to an aversive cue during a continuous performance task. PLoS One 2010;5:e15093.

[61] Thompson JM, Neugebauer V. Amygdala plasticity and pain. Pain Res Manag 2017.

[62] Tillisch K, Mayer EA, Labus JS. Quantitative meta-analysis identifies brain regions activated during rectal distension in irritable bowel syndrome. Gastroenterol 2011;140:91–100.

[63] Walker L, Greene J. The functional disability inventory: measuring a neglected dimension of child health status. J Pediatr Psychol 1991;16:39–58.

[64] Walker LS, Garber J, Greene JW. Psychosocial correlates of recurrent childhood pain: a comparison of pediatric patients with recurrent abdominal pain, organic illness, and psychiatric disorders. J Abnorm Psychol 1993;102:248–258.

[65] Walker LS, Williams SE, Smith CA, Garber J, Van Slyke DA, Lipani T, Greene JW, Mertz H, Naliboff BD. Validation of a symptom provocation test for laboratory studies of abdominal pain and discomfort in children and adolescents. J Pediatr Psychol 2006;31:703–713.

[66] Wang J, Alsop DC, Li L, Listerud J, Gonzalez-At JB, Schnall MD, Detre JA. Comparison of quantitative perfusion imaging using arterial spin labeling at 1.5 and 4.0 Tesla. Magn Reson Med 2002;48:242–254.

[67] Williams SE, Blount RL, Walker LS. Children’s pain threat appraisal and catastrophizing moderate the impact of parent verbal behavior on children’s symptom complaints. J Pediatr Psychol 2011;36:55–63.

[68] Winkler AM, Ridgway GR, Webster MA, Smith SM, Nichols TE. Permutation inference for the general linear model. Neuroimage 2014;92:381–397.

[69] Woolrich MW, Behrens TE, Beckmann CF, Jenkinson M, Smith SM. Multilevel linear modelling for FMRI group analysis using Bayesian inference. Neuroimage 2004;21:1732–1747.

[70] Worsley KJ. Statistical analysis of activation images. Ch 14, in Functional MRI: An Introduction to Methods, eds P Jezzard, PM Matthews and SM Smith Oup, 2001.

[71] Worsley KJ, Evans AC, Marrett S, Neelin P. A three-dimensional statistical analysis for CBF activation studies in human brain. J Cereb Blood Flow Metab 1992;12:900–918.

[72] Zeidan F, Martucci KT, Kraft RA, Gordon NS, McHaffie JG, Coghill RC. Brain mechanisms supporting the modulation of pain by mindfulness meditation. J Neurosci 2011;31:5540–5548.

[73] Zeidan F, Martucci KT, Kraft RA, McHaffie JG, Coghill RC. Neural correlates of mindfulness meditation-related anxiety relief. Soc Cogn Affect Neurosci 2014;9:751–759.

