## Supplementary material for "Changes in amygdalar functional connectivity following the water load symptom provocation task (WL-SPT) in youth with functional abdominal pain disorders": Table 1

**Table 1. Comparison of Youth with FAPD and Healthy Controls**

|  | **FAPD (*n* = 25)**  ***M* (*SD or %*)** | **HC (*n* = 20)**  ***M* (*SD or %*)** |
| --- | --- | --- |
| **Demographics (range)**  Age in years (11-17)* | 13.4 (2.2) | 14.7 (1.8) |
| Sex* |  |  |
| Female | 13 (52%) | 17 (85%) |
| Male | 12 (48%) | 3 (15%) |
| Race |  |  |
| Caucasian/White | 23 (92%) | 16 (80%) |
| Biracial/Other | 2 (8%) | 4 (20%) |
| FDI (0-60)* | 16.5 (12.8) | 0.9 (2.0) |
| Pre-WL-SPT Pain (0-10) |  |  |
| Mean Pain Unpleasantness* | 1.7 (1.9) | 0.0 (0.0) |
| Time 1 | 1.6 (1.8) |  |
| Time 2 | 1.7 (2.0) |  |
| Time 3 | 1.8 (2.2) |  |
| Times 4 | 1.8 (1.8) |  |
| Mean Pain Intensity* | 1.8 (2.1) | 0.0 (0.0) |
| Time 1 | 1.6 (1.8) |  |
| Time 2 | 1.7 (2.0) |  |
| Time 3 | 1.9 (2.3) |  |
| Time 4 | 1.9 (2.3) |  |
| Post-WL-SPT Pain (0-10) |  |  |
| Mean Pain Unpleasantness | 2.5 (2.2) | N/A |
| Time 1 | 2.6 (2.3) |  |
| Time 2 | 2.5 (2.1) |  |
| Time 3 | 2.5 (2.0) |  |
| Time 4 | 2.6 (2.3) |  |
| Mean Pain Intensity | 2.3 (2.3) | N/A |
| Time 1 | 2.3 (2.3) |  |
| Time 2 | 2.2 (2.1) |  |
| Time 3 | 2.3 (2.2) |  |
| Time 4 | 2.4 (2.4) |  |

Note. FAPD- functional abdominal pain disorders; HC- healthy controls; age range of FAPD participants was between 11-16 years and age range of HC was between 12-17 years; FDI-Functional Disability Inventory; WL-SPT- water load symptom provocation task; Mean pain intensity and unpleasantness was calculated using data collected at the four respective time points (e.g., data from Time 1- Time 4); *Significant group difference in gender (X2 = 5.45, p = .020), age (t(43) = 2.18, p < .035), FDI scores (t(43) = 6.00, p < .001), average baseline pain unpleasantness during resting-state scans (t(43) = 4.47, p < .001), and average baseline pain intensity during resting-state scans (t(43) = 4.29, p < .001).
