## Supplementary material for "Changes in amygdalar functional connectivity following the water load symptom provocation task (WL-SPT) in youth with functional abdominal pain disorders": Table 2

| Table 2. FAPD Participants versus HC | | | | | | |
| --- | --- | --- | --- | --- | --- | --- |
| **LEFT AMY** | | | | | | |
| **FAPD > HC** |  |  |  |  |  |  |
| **Brain Structure** | **Cluster Index** | **Z** | **x** | **y** | **z** | **Cluster P value (cluster size)** |
| Lateral Occipital Cortex | 3 | 3.6 | -32 | -76 | 42 | <0.001 (970) |
|  | 3 | 3.57 | -30 | -76 | 38 | <0.001 (970) |
|  | 3 | 3.51 | -28 | -66 | 36 | <0.001 (970) |
|  | 3 | 3.48 | -26 | -62 | 34 | <0.001 (970) |
|  | 3 | 3.21 | -34 | -76 | 26 | <0.001 (970) |
|  | 3 | 3.18 | -20 | -64 | 34 | <0.001 (970) |
| Right Cerebral Cortex White Matter | 2 | 3.17 | 16 | 22 | 44 | 0.002 (757) |
| Superior Frontal Gyrus | 2 | 3.17 | 22 | 20 | 42 | 0.002 (757) |
| Middle Frontal Gyrus | 2 | 3.15 | 26 | 4 | 50 | 0.002 (757) |
|  | 2 | 3.12 | 36 | 16 | 44 | 0.002 (757) |
| Superior Frontal Gyrus | 2 | 3.06 | 20 | 22 | 54 | 0.002 (757) |
| Middle Frontal Gyrus | 2 | 3.05 | 28 | 12 | 56 | 0.002 (757) |
| Lateral Occipital Cortex | 1 | 3.58 | 40 | -58 | 42 | 0.010 (566) |
|  | 1 | 3.33 | 34 | -66 | 48 | 0.010 (566) |
|  | 1 | 3.12 | 38 | -68 | 42 | 0.010 (566) |
|  | 1 | 3.02 | 40 | -58 | 36 | 0.010 (566) |
| Angular Gyrus | 1 | 3.02 | 42 | -54 | 52 | 0.010 (566) |
|  | 1 | 2.98 | 40 | -56 | 48 | 0.010 (566) |

| **RIGHT AMY** |  |  |  |  |  |  |
| --- | --- | --- | --- | --- | --- | --- |
| **HC > FAPD** |  |  |  |  |  |  |
| **Brain Structure** | **Cluster Index** | **Z** | **x** | **y** | **z** | **Cluster P value (cluster size)** |
| Occipital Pole | 2 | 3.3 | -36 | -98 | 12 | 0.010 (588) |
|  | 2 | 3.16 | -32 | -98 | 12 | 0.010 (588) |
|  | 2 | 3.12 | -26 | -90 | 10 | 0.010 (588) |
| Left Cerebral Cortex | 2 | 3.07 | -42 | -92 | 18 | 0.010 (588) |
| Lateral Occipital Cortex | 2 | 3.02 | -44 | -88 | 0 | 0.010 (588) |
|  | 2 | 2.9 | -50 | -78 | 16 | 0.010 (588) |
|  | 1 | 3.64 | 44 | -80 | 2 | 0.011 (577) |
| Occipital Pole | 1 | 3.59 | 28 | -96 | 12 | 0.011 (577) |
|  | 1 | 3.06 | 38 | -92 | 6 | 0.011 (577) |
| Occipital Fusiform Gyrus | 1 | 3.03 | 28 | -72 | 0 | 0.011 (577) |
|  | 1 | 3 | 30 | -68 | 0 | 0.011 (577) |
| Lateral Occipital Cortex | 1 | 2.98 | 40 | -62 | 0 | 0.011 (577) |

| **FAPD > HC** |  |  |  |  |  |  |
| --- | --- | --- | --- | --- | --- | --- |
| **Brain Structure** | **Cluster Index** | **Z** | **x** | **y** | **z** | **Cluster P value (cluster size)** |
| Lateral Occipital Cortex | 3 | 3.5 | -46 | -74 | 32 | 0.001 (803) |
|  | 3 | 3.26 | -18 | -68 | 38 | 0.001 (803) |
|  | 3 | 3.19 | -32 | -82 | 36 | 0.001 (803) |
| Precuneous Cortex | 3 | 3.18 | -20 | -64 | 30 | 0.001 (803) |
| Lateral Occipital Cortex | 3 | 3.15 | -36 | -74 | 36 | 0.001 (803) |
|  | 3 | 3.15 | -30 | -64 | 34 | 0.001 (803) |
| Precuneous Cortex | 2 | 3.64 | 18 | -50 | 16 | 0.008 (610) |
|  | 2 | 3.29 | 24 | -56 | 16 | 0.008 (610) |
| Supracalcarine Cortex | 2 | 3.28 | 26 | -60 | 16 | 0.008 (610) |
| Precuneous Cortex | 2 | 3.24 | 20 | -60 | 14 | 0.008 (610) |
|  | 2 | 3.15 | 12 | -62 | 28 | 0.008 (610) |
|  | 2 | 2.96 | 16 | -62 | 24 | 0.008 (610) |
|  | 1 | 3.7 | -20 | -58 | 18 | 0.014 (555) |
| Cingulate Gyrus | 1 | 3.32 | -8 | -42 | 24 | 0.014 (555) |
|  | 1 | 3.32 | -8 | -48 | 16 | 0.014 (555) |
|  | 1 | 3.13 | -14 | -48 | 30 | 0.014 (555) |
| Precuneous Cortex | 1 | 3.11 | -4 | -58 | 8 | 0.014 (555) |
|  | 1 | 2.93 | -6 | -66 | 22 | 0.014 (555) |

*Note.* FAPD – functional abdominal pain disorders; HC – healthy controls; AMY - amygdala.
