## Supplementary figures and images for "Changes in amygdalar functional connectivity following the water load symptom provocation task (WL-SPT) in youth with functional abdominal pain disorders"

### Figure 2

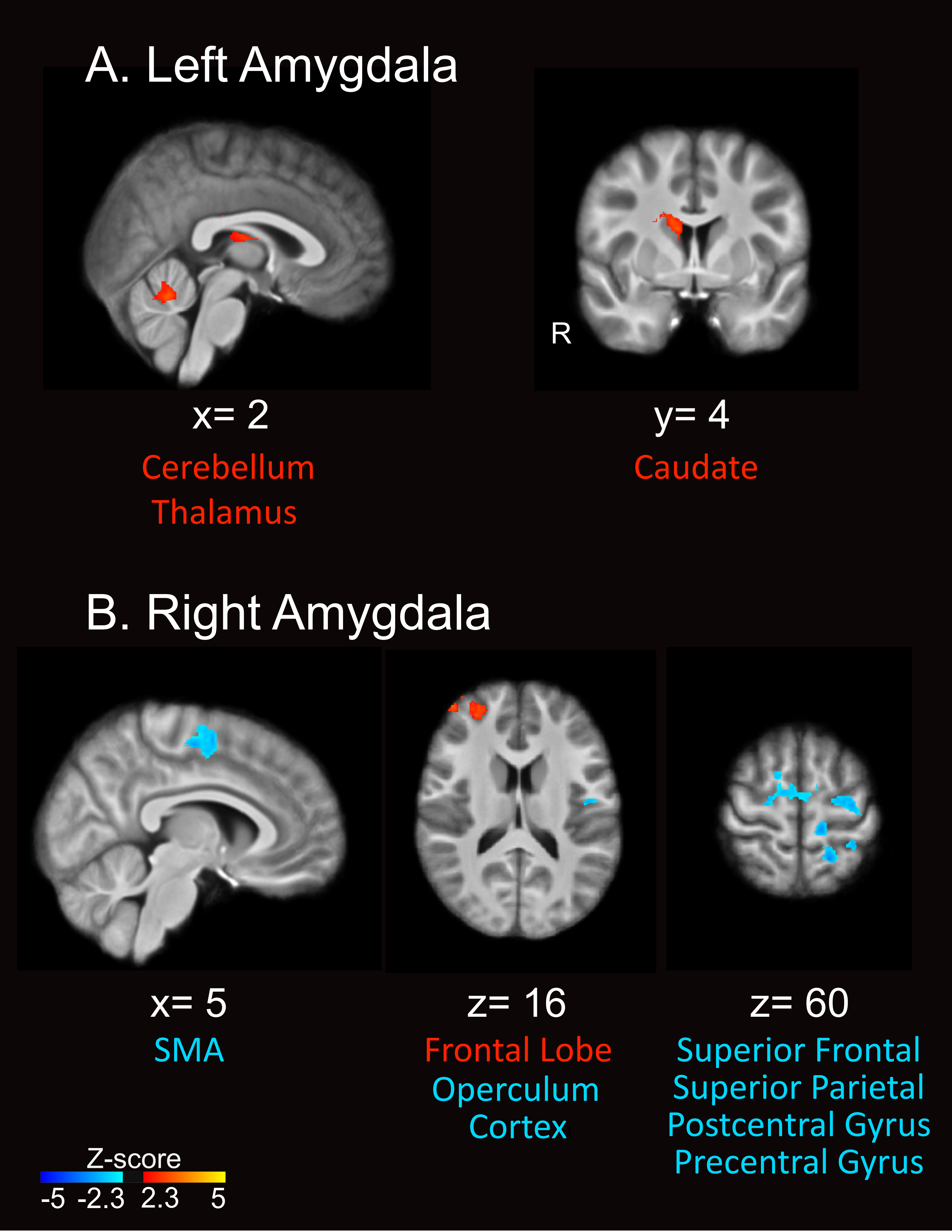

### Figure 3

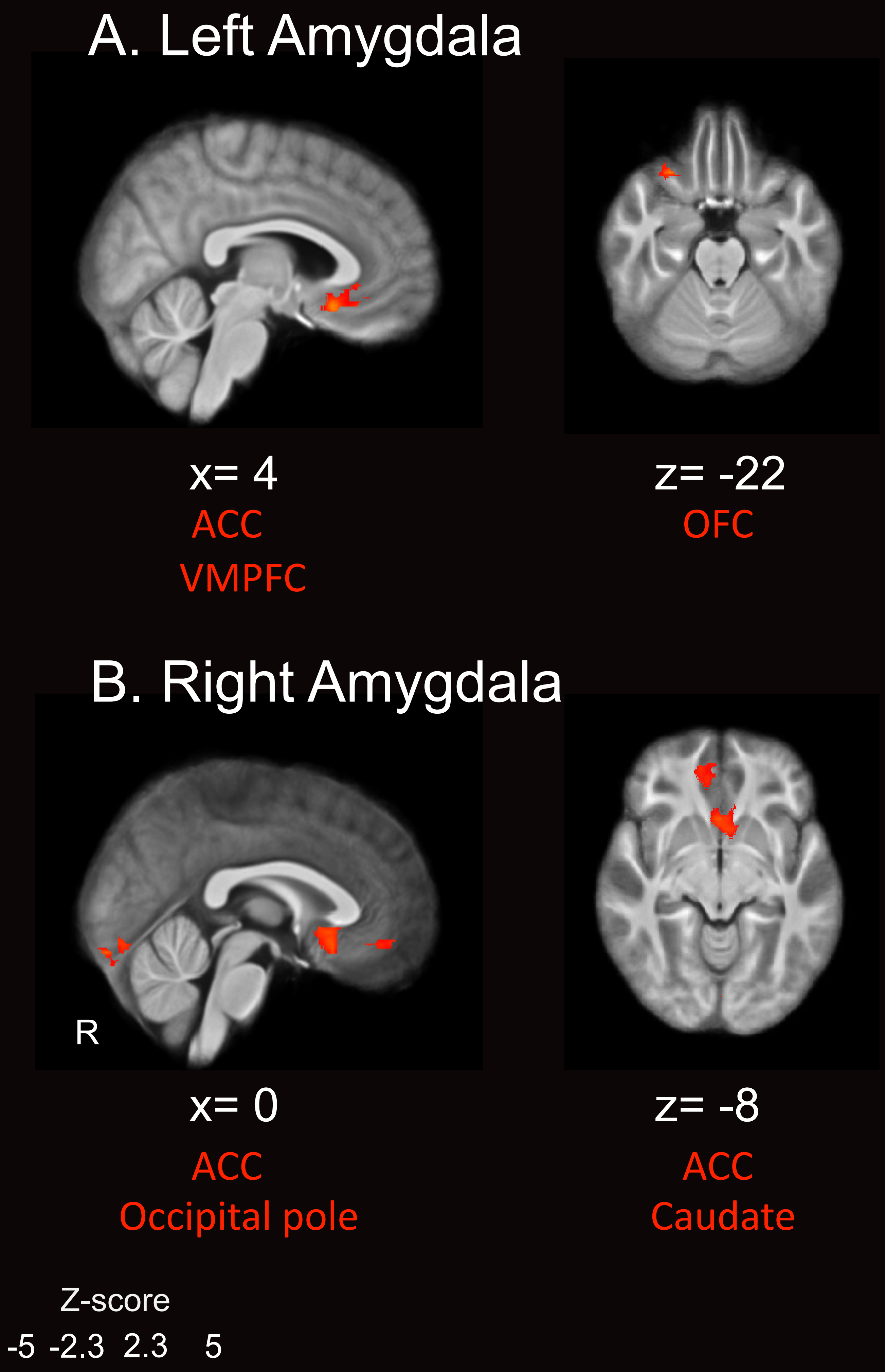

### Figure 4

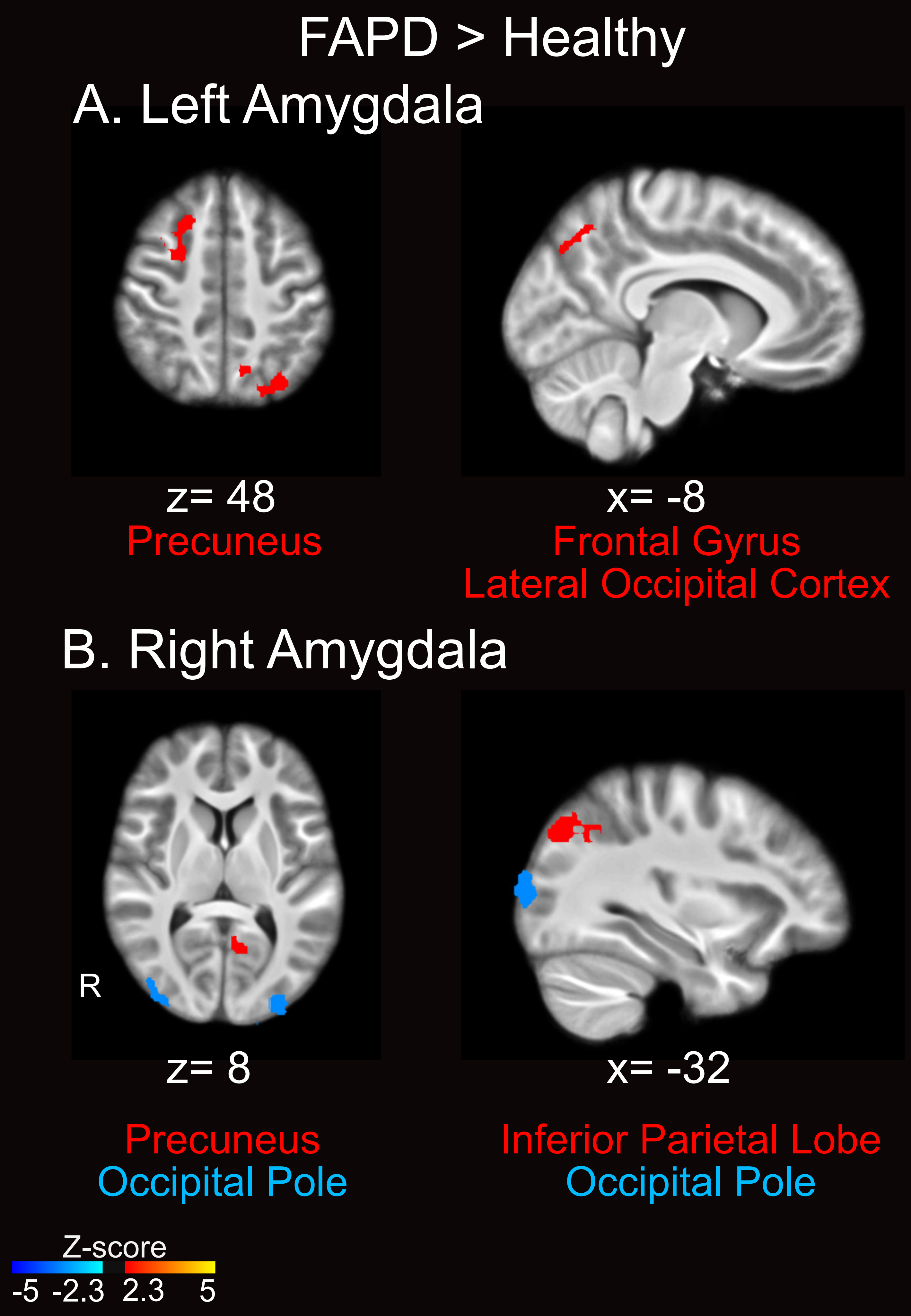
