## Supplementary material for "Changes in amygdalar functional connectivity following the water load symptom provocation task (WL-SPT) in youth with functional abdominal pain disorders": Figure 1

Mean Pain Intensity (0-10)

| Pre WL-SPT Post WL-SPT | Intensity |
| --- | --- |
| T1 | 1.612 |
| T2 | 1.708 |
| T3 | 1.888 |
| T4 | 1.944 |
| T1 | 2.292 |
| T2 | 2.176 |
| T3 | 2.312 |
| T4 | 2.398 |

Mean Pain Unpleasantness (0-10)

| Pre WL-SPT Post WL-SPT | Unpleasantness |
| --- | --- |
| T1 | 1.56 |
| T2 | 1.716 |
| T3 | 1.784 |
| T4 | 1.82 |
| T1 | 2.632 |
| T2 | 2.456 |
| T3 | 2.526 |
| T4 | 2.618 |

*Note.* WL-SPT - Water loading symptom provocation task; T – time; (total possible range). Two-way ANOVAs revealed significant differences by condition (pre versus post WL-SPT) on pain unpleasantness (omnibus, F (1, 24) = 20.241, p < .001; main effect, p < 0.001) and pain intensity (omnibus, F (1, 24) = 7.215, p = .013; main effect, p = 0.013) only. There was no significant differences by time, nor was there a time by condition interaction (all p’s > 0.05).
